# Socioeconomic Inequalities and Environmental Determinants of Child Undernutrition in Cambodia: An Analysis of the 2021–22 Demographic and Health Survey

**DOI:** 10.64898/2026.05.06.26352583

**Authors:** Samnang Um, Leng Dany, Sok Sakha, Pichsokkim Pav, Channnarong Phan, Pall Chamroen, Chantrea Sieng, Sovandara Heng

**Affiliations:** Center for Health Research and Policy Support, National Institute of Public Health, Phnom Penh, Cambodia; Department of Pediatrics, Elite Angkor Clinic, Siem Reap, Cambodia; Department of Laboratory, Calmette Hospital, Phnom Penh, Cambodia; Health Poverty Action, Phnom Penh, Cambodia; School of Public Health, National Institute of Public Health, Phnom Penh, Cambodia; Department of Orthodontics, Faculty of Dentistry, University of Puthisastra, Phnom Penh, Cambodia; Department of Outpatient, Calmette Hospital, Phnom Penh, Cambodia; Department of Mental Health and Substance Abuse, Khmer Soviet Friendship Hospital, Phnom Penh, Cambodia

**Keywords:** Child undernutrition, stunting, wasting, underweight, under-five children, Cambodia, socioeconomic inequality, water sanitation and hygiene, CDHS

## Abstract

Childhood undernutrition remains a major public health problem in low- and middle-income countries, including Cambodia, despite improvements in economic development and health services. To better understand child undernutrition in Cambodia, we examined the prevalence, socio-economic and environmental determinants, and associated factors of stunting, wasting, and underweight among children aged 0–59 months using the 2021–22 Cambodia Demographic and Health Survey (CDHS). This study included 3,821 weighted children aged 0–59 months. The prevalence of stunting, wasting, and underweight was 20.4%, 9.4%, and 15.9%, respectively. The highest burden of stunting was observed among children aged 12–23 months. Children from the poorest households consistently contributed the largest proportion of undernutrition cases across all three outcomes. After adjusting for other variables, children aged 12–23 months had higher odds of stunting compared with infants (AOR = 2.64; 95% CI: 1.87–3.74). Male children had increased odds of stunting (AOR = 1.33; 95% CI: 1.10–1.61), wasting (AOR = 1.35; 95% CI: 1.01–1.82), and underweight (AOR = 1.28; 95% CI: 1.02–1.59). Children from richer (AOR = 0.55; 95% CI: 0.31–0.95) and richest households (AOR = 0.33; 95% CI: 0.16–0.68) had lower odds of stunting. Maternal secondary or higher education was associated with lower odds of wasting (AOR = 0.49; 95% CI: 0.28–0.84) and underweight (AOR = 0.51; 95% CI: 0.32–0.79). Access to bottled water and digital connectivity were also protective against undernutrition. Conversely, poor household environmental conditions were associated with increased odds of undernutrition. Interaction analysis showed a stronger protective effect of maternal education among wealthier households, indicating a synergistic effect of socioeconomic advantage. Childhood undernutrition remains highly prevalent in Cambodia. Public health interventions and policies should prioritize improving complementary feeding practices, reducing socioeconomic inequalities, and strengthening multisectoral actions across nutrition, education, WASH, and social protection systems.

## Introduction

Child undernutrition remains a major global public health challenge, particularly in low- and middle-income countries. It is commonly manifested as stunting (low height-for-age), wasting (low weight-for-height), and underweight (low weight-for-age), each defined as a Z-score below −2 standard deviations from the median of the World Health Organization (WHO) Child Growth Standards [1, 2]. These indicators capture distinct but overlapping dimensions of nutritional deprivation, reflecting chronic, acute, and composite forms of malnutrition, respectively. In 2022, an estimated 149 million children under five years were stunted, and 45 million were wasted worldwide [2]. Undernutrition contributes to nearly half of all deaths among children under five and has profound consequences for physical growth, cognitive development, and long-term human capital formation [3].

In Southeast Asia, the burden of child undernutrition remains substantial, with approximately 27.4% of children under five affected by stunting [4, 5]. Cambodia has made notable progress over the past decade; however, undernutrition continues to pose a significant public health concern. The persistence of undernutrition despite economic growth highlights the unequal distribution of health gains and the presence of structural vulnerabilities among disadvantaged populations. According to the 2021–22 Cambodia Demographic and Health Survey (CDHS), the prevalence of stunting, wasting, and underweight remains high at 23.2%, 9.9%, and 16.4%, respectively [6]. These levels exceed regional benchmarks and indicate persistent nutritional vulnerability, particularly among disadvantaged populations.

Despite sustained economic growth and improvements in health service coverage, progress in reducing child undernutrition in Cambodia has been uneven. Marked disparities persist across socioeconomic groups, geographic regions, and environmental conditions, with children in rural and climate-vulnerable settings disproportionately affected [6, 7]. These inequalities reflect the multifactorial nature of undernutrition, which extends beyond immediate dietary intake and disease to include broader social, economic, and environmental determinants [3].

Recent nationally representative studies in Cambodia further highlight the interconnected nature of child health challenges. Evidence from childhood anemia research indicates that anemia remains highly prevalent, affecting more than half of children under five, with strong links to maternal anemia, poor nutritional status, infectious diseases, and household poverty, as well as pronounced geographic disparities across provinces [8]. In contrast, studies on acute respiratory infections (ARI) and childhood diarrhea demonstrate substantial declines over the past two decades, reflecting improvements in healthcare access, immunization, and WASH interventions [9, 10]. However, these conditions continue to disproportionately affect younger children, those exposed to household smoking, and those living in poorer households with inadequate sanitation [9, 10]. These contrasting trends suggest that while Cambodia has made progress in reducing infectious diseases, nutrition-related conditions remain persistent and are deeply rooted in structural inequalities. This shift reflects a broader epidemiological transition, where chronic and nutrition-related conditions increasingly dominate child health outcomes.

The determinants of child undernutrition are well conceptualized within the UNICEF framework, which categorizes causes into immediate (inadequate dietary intake and disease), underlying (household food insecurity, inadequate care and feeding practices, unhealthy environments, and limited health services), and basic factors (socioeconomic conditions, governance, and structural inequities) [3]. This framework provides a critical lens for understanding how distal structural factors interact with proximal biological mechanisms to influence child growth outcomes. In this context, factors such as household wealth, maternal education, housing quality, water and sanitation conditions, and access to information and health services play critical roles in shaping child nutritional outcomes [6, 7].

Cambodia has committed to achieving Sustainable Development Goal (SDG) 2.2, which aims to end all forms of malnutrition by 2030, including reducing stunting and wasting among children under five [2]. National targets further aim to reduce stunting by 40% and wasting to below 5% by 2025 [11, 12]. However, achieving these targets requires a comprehensive understanding of the multilevel drivers of undernutrition within the Cambodian context.

While previous studies have examined selected determinants of child malnutrition in Cambodia, there remains a need for updated, nationally representative evidence that integrates individual, maternal, household, environmental, and geographic factors [7]. Moreover, existing evidence has largely examined child health outcomes in isolation (e.g., anemia, ARI, or diarrhea), with limited integration of these findings into a broader framework of nutritional vulnerability [8–10]. Bridging this gap is essential to better understand how overlapping risk factors interact to shape child undernutrition in Cambodia. Importantly, limited studies have simultaneously examined multiple domains of determinants—ranging from socioeconomic and environmental to digital and health access factors—within a unified analytical framework using recent nationally representative data.

Therefore, this study aims to assess the prevalence and identify the socioeconomic and environmental determinants of stunting, wasting, and underweight among children under five years in Cambodia using data from the 2021–22 CDHS. By incorporating interaction effects and multiple determinant domains, this study advances existing evidence by highlighting not only independent risk factors but also how these factors jointly shape child nutritional outcomes.

## Material and Methods

### Ethical Consideration

The original data collection tools and protocols for the 2021–22 Cambodia Demographic and Health Survey (CDHS) were reviewed and approved by the National Ethics Committee for Health Research (NECHR) in Cambodia on May 10, 2021 (Ref #083 NECHR), and the Institutional Review Board (IRB) of ICF in Rockville, Maryland, USA. Prior to the commencement of fieldwork, written informed consent was obtained from all participants; for children under the age of 18, consent was obtained from their parents or legal caregivers [6].

This study involved a secondary analysis of the 2021–22 CDHS dataset. Access to the raw data was granted by the DHS Program following a formal request. Because the dataset is fully de-identified and publicly available for research purposes, this study was exempt from further institutional ethical review. The data were used in strict accordance with the terms and conditions prescribed by The DHS Program [13].

### Data Source and Study Design

Data were derived from the 2021–22 Cambodia Demographic and Health Survey (CDHS), a nationally representative cross-sectional survey implemented by the National Institute of Statistics (NIS) in collaboration with the Ministry of Health (MoH). The survey utilized a two-stage stratified cluster sampling design. In the first stage, 709 enumeration areas (clusters), comprising 241 urban and 468 rural areas, were selected with probability proportional to size. In the second stage, a systematic sample of households was drawn from each cluster, totaling 21,270 selected households [6].

Fieldwork was conducted between September 15, 2021, and February 15, 2022. Of the 20,967 occupied households identified, 20,806 were successfully interviewed, representing a household response rate of 99%. From these households, 19,496 women aged 15–49 were successfully interviewed, yielding a 98% response rate. To analyze the determinants of childhood undernutrition, the Children’s Recode (KR), Women’s Recode (IR), and Household Recode (HR) files were merged using unique cluster, household, and individual identifiers to create a comprehensive multi-level dataset [6].

### Study population and analytical sample

The initial study population included all children aged 0–59 months identified across the 20,806 interviewed households (N = 8,153). However, in accordance with the 2021–22 CDHS protocol, anthropometric data collection was restricted to a 50% subsample of households—specifically those selected for the men’s survey [6]. As confirmed by the survey records, 4,018 children (49.3% of the total children identified) resided in these designated households and were successfully measured for height and weight. To ensure the technical integrity of the results, these records were screened for data quality. In this study, the final analytical sample of 4,018 children represents all those with valid anthropometric measurements within the subsample. This sample was then weighted using the survey’s sampling weights to ensure the results are nationally representative of all children under five in Cambodia.

### Measurement of Variables Outcome Variables

Childhood undernutrition was assessed based on the 2006 World Health Organization (WHO) Child Growth Standards [1]. Three primary anthropometric indices were utilized as binary dependent variables: 1) **Stunting:** Defined as a height-for-age Z-score (HAZ) < −2 standard deviations (SD) from the median of the reference population (coded: 1 = stunted, 0 = not stunted). 2) **Wasting:** Defined as a weight-for-height Z-score (WHZ) < −2 SD from the median (coded: 1 = wasted, 0 = not wasted). 3) **Underweight**: Defined as a weight-for-age Z-score (WAZ) < −2 SD from the median (coded: 1 = underweight, 0 = not underweight).

### Independent Variables

A total of five conceptual domains were constructed based on literature regarding the determinants of malnutrition in low- and middle-income settings:

### Child and Maternal Characteristics

Child-level variables included sex (male, female) and age group (0–11, 12–23, 24–35, 36–47, and 48–59 months), Recent diarrhea and fever in the past 2 weeks (yes vs no). Maternal characteristics included age (15–19, 20–29, 30–39, and 40–49 years), marital status (currently married/cohabiting vs. not married), educational attainment (no education, primary, secondary, or higher), and occupational status (not working, professional/managerial, clerical, sales/services, agricultural, and domestic/manual labor).

### Household Socioeconomic Status (SES)

SES was captured through the wealth index quintiles (poorest, poorer, middle, richer, and richest) [6, 14, 15], household size (≤3, 4–6, and ≥7 members). Productive assets were measured by agricultural land ownership (no land, 1–5, 5–20, and >20 hectares). Housing quality was a composite variable based on improved vs. unimproved floor, wall, and roof materials[14]. Household crowding was defined as >3 persons per sleeping room [14].

### Water, Sanitation, and Hygiene (WASH) and Environment

WASH variables were classified according to the WHO/UNICEF Joint Monitoring Programme (JMP) standards [16]. These included drinking water source (improved, bottled, unimproved), water treatment (yes/no), sanitation facility type (improved, unimproved, open defecation), toilet sharing (private vs. shared), and handwashing ready (availability of a station with both water and soap). Additionally, cooking energy was dichotomized into clean vs. polluting fuels, and household smoking status was recorded.

### Health Access and Media Exposure

Health-related factors included travel time to the nearest health facility (≤15, 16–30, and >30 minutes) and health insurance coverage. Media exposure was categorized by the number of sources (newspaper, radio, TV) accessed. Digital connectivity was measured through mobile phone type (none, basic, smartphone) and frequency of internet use (never, less than weekly, daily).

### Geographic Factors

To account for spatial variations, place of residence (urban vs. rural) and geographic region were included. The 25 provinces were recoded into five established ecological zones: Phnom Penh, Plains, Tonle Sap, Coastal, and Mountainous regions [6].

### Statistical analysis

We analyzed the data from the 2021–22 Cambodia Demographic and Health Survey (CDHS) using Stata/MP version 18.0. To ensure the results represent the national population, we accounted for the complex survey design by incorporating sampling weights (v005/1,000,000), primary sampling units (v021), and strata (v023) through the svyset command. Our initial work involved data cleaning, variable labeling, and constructing the key outcome and explanatory variables. We used descriptive statistics and missing data assessments to understand the distribution of the study population and calculated the weighted prevalence of stunting, wasting, and underweight with 95% confidence intervals.

### Bivariate Analysis

We first conducted bivariate analyses using survey-adjusted cross-tabulations (svy: tab) to look for associations between each independent variable and the three nutritional outcomes. We used design-based Pearson chi-square tests to evaluate statistical significance. The variables we examined included child characteristics (age, sex, and recent diarrhea or fever), maternal factors (age, education, occupation, and marital status), and household socioeconomic indicators (wealth index, size, and number of children under five). We also looked at housing conditions, land ownership, WASH factors (water source, treatment, and sanitation), household assets, smoking status, and access to health services and media.

### Multivariable Modeling

Any variable with a p-value < 0.10 in the bivariate stage was included in the multivariable logistic regression models [17, 18]. This threshold was selected to avoid premature exclusion of potentially important predictors in multivariable analysis. We fitted three separate survey-weighted models for stunting, wasting, and underweight using the svy: logistic command. We reported adjusted odds ratios (AORs) with 95% confidence intervals to identify the factors independently linked to undernutrition, with statistical significance set at p < 0.05.

### Model Diagnostics and Robustness

To enhance model validity, multiple diagnostic procedures were conducted, including goodness-of-fit testing, multicollinearity assessment, and discrimination analysis.

### Interaction Effects

Interaction terms were introduced to assess effect modification and to explore whether the relationship between key predictors and undernutrition outcomes varied across population subgroups.

## Results

### Description of the study population

The final analytical sample comprised 4,018 children aged 0–59 months, representing a weighted population of 3,821. The sample was nearly evenly split by sex (51.2% male) and age group, with the largest proportion aged 12–23 months (21.5%). Most mothers were aged 20–39 years (90.5%), currently married (93.8%), and employed (62.3%). Regarding maternal education, 48.5% had completed secondary or higher education, while 10.4% had no formal schooling.

Household wealth was distributed relatively evenly across quintiles, though the largest share belonged to the poorest group (22.9%). While most households were of medium size (67.7%), many faced challenging living conditions: 57.5% lived in unimproved housing, 56.3% lived in crowded environments, and 33.6% reported exposure to household smoking.

In terms of infrastructure and WASH (Water, Sanitation, and Hygiene), access to electricity was high (90.8%), and 63.0% utilized improved drinking water. However, significant gaps remained, with 11.3% practicing open defecation and 30.5% not treating their drinking water. Although smartphone ownership was prevalent (76.1%), 38.2% of households never used the internet and 55.6% had no exposure to traditional media. Geographically, 61.6% of the population resided in rural areas (**Table 1**). These findings indicate that a substantial proportion of Cambodian children are exposed to structural and environmental risk factors that may adversely affect nutritional outcomes.

**Table 1.**
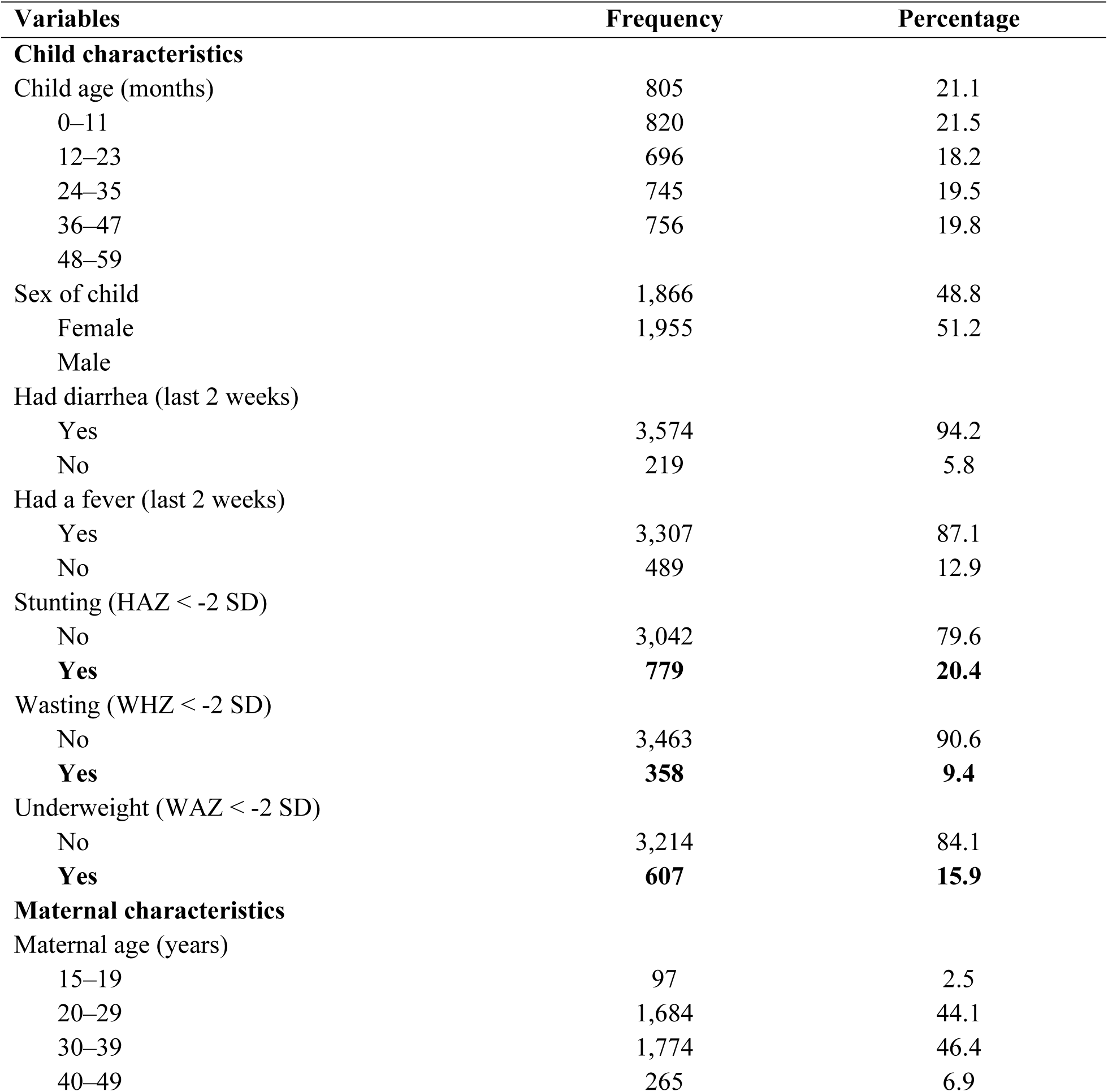
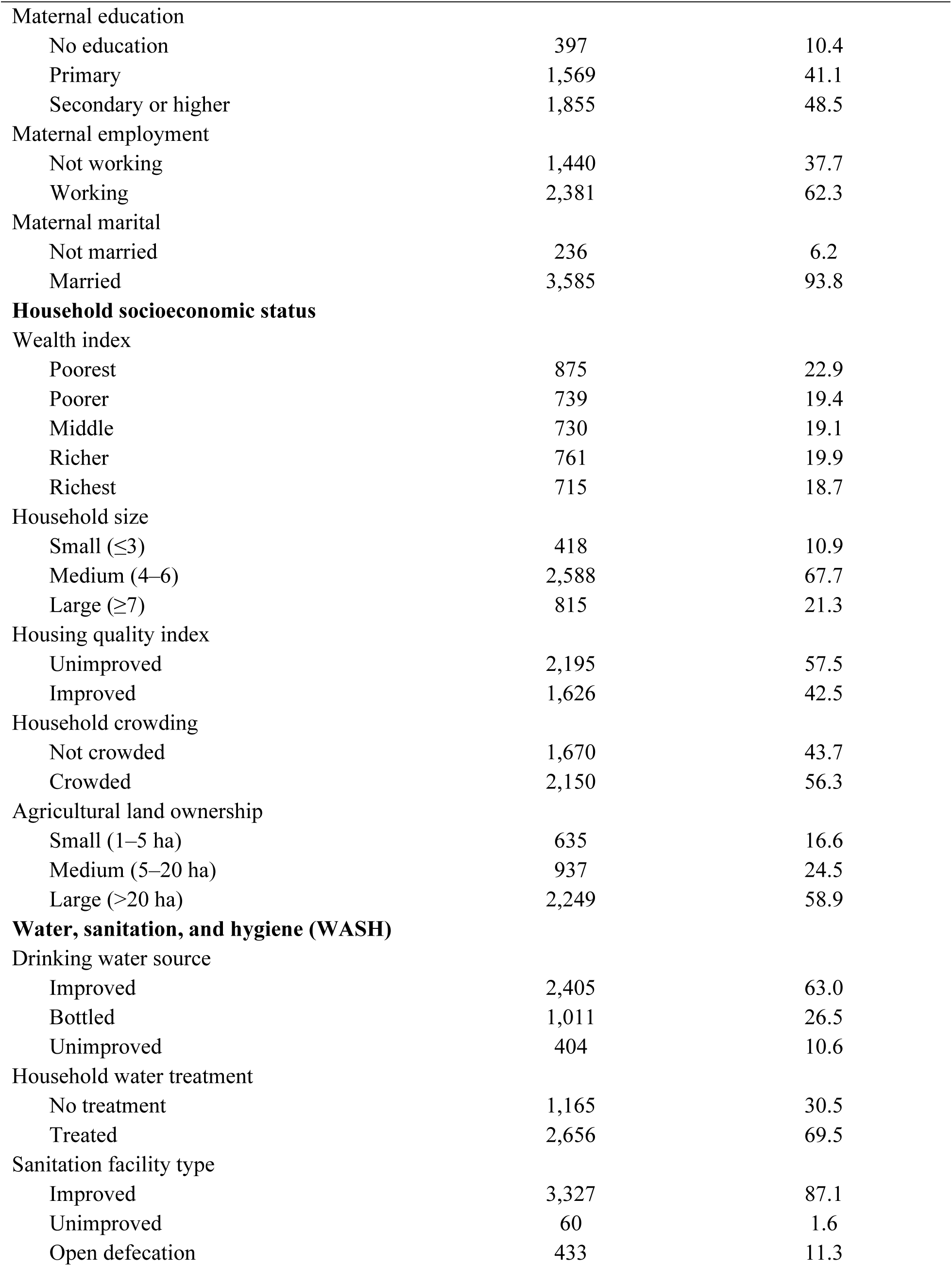
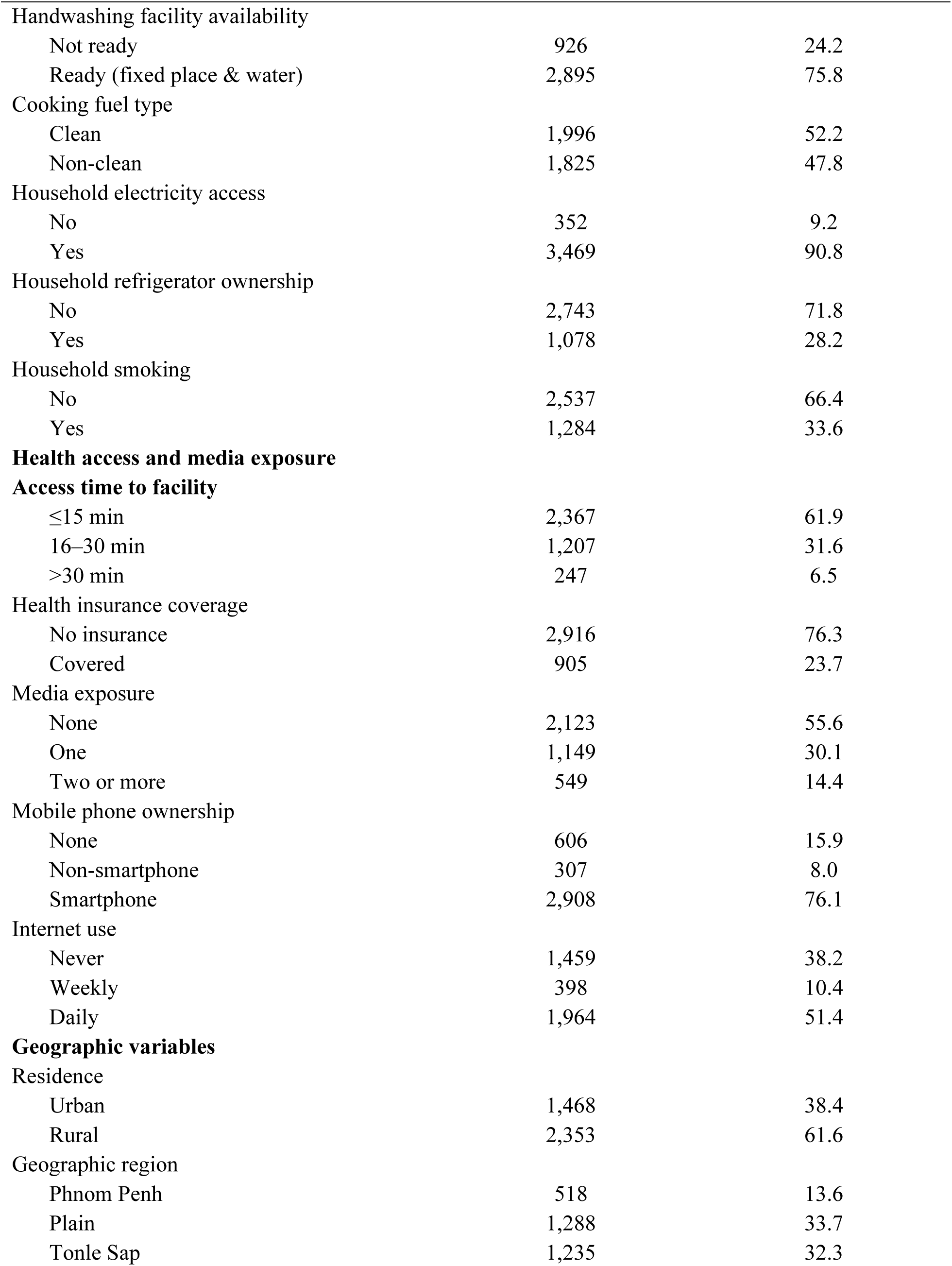
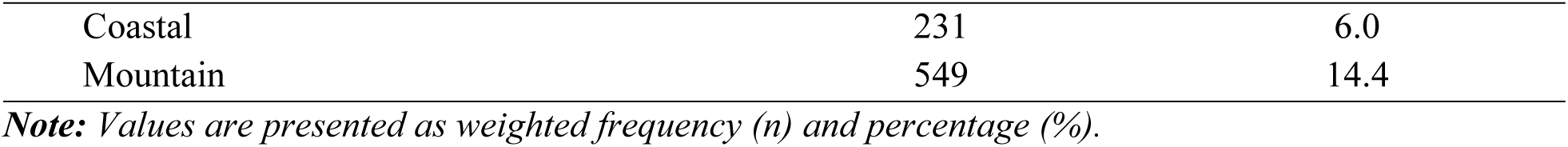
Descriptive of children aged 0–59 months in Cambodia (CDHS 2021–2022, N = 3,821)

| Variables | Frequency | Percentage |
| --- | --- | --- |
| **Child characteristics** |  |  |
| Child age (months) | 805 | 21.1 |
| 0–11 | 820 | 21.5 |
| 12–23 | 696 | 18.2 |
| 24–35 | 745 | 19.5 |
| 36–47 | 756 | 19.8 |
| 48–59 |  |  |
| Sex of child | 1,866 | 48.8 |
| Female | 1,955 | 51.2 |
| Male |  |  |
| Had diarrhea (last 2 weeks) |  |  |
| Yes | 3,574 | 94.2 |
| No | 219 | 5.8 |
| Had a fever (last 2 weeks) |  |  |
| Yes | 3,307 | 87.1 |
| No | 489 | 12.9 |
| Stunting (HAZ < -2 SD) |  |  |
| No | 3,042 | 79.6 |
| **Yes** | **779** | **20.4** |
| Wasting (WHZ < -2 SD) |  |  |
| No | 3,463 | 90.6 |
| **Yes** | **358** | **9.4** |
| Underweight (WAZ < -2 SD) |  |  |
| No | 3,214 | 84.1 |
| **Yes** | **607** | **15.9** |
| **Maternal characteristics** |  |  |
| Maternal age (years) |  |  |
| 15–19 | 97 | 2.5 |
| 20–29 | 1,684 | 44.1 |
| 30–39 | 1,774 | 46.4 |
| 40–49 | 265 | 6.9 |
| Maternal education |  |  |
| No education | 397 | 10.4 |
| Primary | 1,569 | 41.1 |
| Secondary or higher | 1,855 | 48.5 |
| Maternal employment |  |  |
| Not working | 1,440 | 37.7 |
| Working | 2,381 | 62.3 |
| Maternal marital |  |  |
| Not married | 236 | 6.2 |
| Married | 3,585 | 93.8 |
| **Household socioeconomic status** |  |  |
| Wealth index |  |  |
| Poorest | 875 | 22.9 |
| Poorer | 739 | 19.4 |
| Middle | 730 | 19.1 |
| Richer | 761 | 19.9 |
| Richest | 715 | 18.7 |
| Household size |  |  |
| Small ($\leq 3$) | 418 | 10.9 |
| Medium (4–6) | 2,588 | 67.7 |
| Large ($\geq 7$) | 815 | 21.3 |
| Housing quality index |  |  |
| Unimproved | 2,195 | 57.5 |
| Improved | 1,626 | 42.5 |
| Household crowding |  |  |
| Not crowded | 1,670 | 43.7 |
| Crowded | 2,150 | 56.3 |
| Agricultural land ownership |  |  |
| Small (1–5 ha) | 635 | 16.6 |
| Medium (5–20 ha) | 937 | 24.5 |
| Large ($> 20$ ha) | 2,249 | 58.9 |
| **Water, sanitation, and hygiene (WASH)** |  |  |
| Drinking water source |  |  |
| Improved | 2,405 | 63.0 |
| Bottled | 1,011 | 26.5 |
| Unimproved | 404 | 10.6 |
| Household water treatment |  |  |
| No treatment | 1,165 | 30.5 |
| Treated | 2,656 | 69.5 |
| Sanitation facility type |  |  |
| Improved | 3,327 | 87.1 |
| Unimproved | 60 | 1.6 |
| Open defecation | 433 | 11.3 |

| **Variables** | **Frequency** | **Percentage** |
| --- | --- | --- |
| Handwashing facility availability |  |  |
| Not ready | 926 | 24.2 |
| Ready (fixed place & water) | 2,895 | 75.8 |
| Cooking fuel type |  |  |
| Clean | 1,996 | 52.2 |
| Non-clean | 1,825 | 47.8 |
| Household electricity access |  |  |
| No | 352 | 9.2 |
| Yes | 3,469 | 90.8 |
| Household refrigerator ownership |  |  |
| No | 2,743 | 71.8 |
| Yes | 1,078 | 28.2 |
| Household smoking |  |  |
| No | 2,537 | 66.4 |
| Yes | 1,284 | 33.6 |
| **Health access and media exposure** |  |  |
| **Access time to facility** |  |  |
| ≤15 min | 2,367 | 61.9 |
| 16–30 min | 1,207 | 31.6 |
| >30 min | 247 | 6.5 |
| Health insurance coverage |  |  |
| No insurance | 2,916 | 76.3 |
| Covered | 905 | 23.7 |
| Media exposure |  |  |
| None | 2,123 | 55.6 |
| One | 1,149 | 30.1 |
| Two or more | 549 | 14.4 |
| Mobile phone ownership |  |  |
| None | 606 | 15.9 |
| Non-smartphone | 307 | 8.0 |
| Smartphone | 2,908 | 76.1 |
| Internet use |  |  |
| Never | 1,459 | 38.2 |
| Weekly | 398 | 10.4 |
| Daily | 1,964 | 51.4 |
| **Geographic variables** |  |  |
| Residence |  |  |
| Urban | 1,468 | 38.4 |
| Rural | 2,353 | 61.6 |
| Geographic region |  |  |
| Phnom Penh | 518 | 13.6 |
| Plain | 1,288 | 33.7 |
| Tonle Sap | 1,235 | 32.3 |
| Coastal | 231 | 6.0 |
| Mountain | 549 | 14.4 |

*Note: Values are presented as weighted frequency (n) and percentage (%).*

### Prevalence of Undernutrition and Childhood Illness

The weighted prevalence of undernutrition among Cambodian children remains significant. Stunting affected 20.4% (95% CI: 18.5–22.2) of the population, while 9.4% (95% CI: 8.1–10.6) were wasted and 15.9% (95% CI: 14.4–17.4) were underweight (**Table 1**). Regarding recent morbidity, 5.8% of children had experienced diarrhea, and 12.9% had a fever in the two weeks preceding the survey, highlighting a persistent burden of acute illness. The coexistence of undernutrition and infectious morbidity highlights the dual burden of disease among children in Cambodia.

### Prevalence of stunting, wasting, and underweight by background characteristics

**Table 2** presents the distribution of undernutrition across various socio-demographic and environmental factors.

**Table 2.** Weighted prevalence of stunting, wasting, and underweight by socio-demographic, maternal, household, and environmental factors among children under five in Cambodia (DHS 2021–2022), (N = 3,821 weighted)

| Variables | Stunting<br>n (%) | p-value | Wasting<br>n(%) | p-value | Underweight<br>n(%) | p-value |
| --- | --- | --- | --- | --- | --- | --- |
| **Child characteristics** |  |  |  |  |  |  |
| Child age (months) |  |  |  |  |  |  |
| 0–11 | 95.9 (12.3) | <0.001 | 94.1 (26.3) | 0.240 | 94.1 (26.3) | 0.240 |
| 12–23 | 218 (28.0) |  | 57.1 (16.0) |  | 57.1 (16.0) |  |
| 24–35 | 153 (19.7) |  | 66.0 (18.5) |  | 66.0 (18.5) |  |
| 36–47 | 156 (20.1) |  | 68.5 (19.1) |  | 68.5 (19.1) |  |
| 48–59 | 156 (20.0) |  | 71.9 (20.1) |  | 71.9 (20.1) |  |
| Sex of child |  |  |  |  |  |  |
| Female | 337 (43.2) | 0.002 | 150 (41.8) | 0.030 | 150 (41.8) | 0.030 |
| Male | 442 (56.8) |  | 208 (58.2) |  | 208 (58.2) |  |
| Had diarrhea (last 2 weeks) |  |  |  |  |  |  |
| No | 725 (20.3) | 0.193 | 334 (9.35) | 0.566 | 574 (16.1) | 0.631 |
| Yes | 54 (24.4) |  | 24 (10.7) |  | 33 (14.9) |  |
| Had a fever (last 2 weeks) |  |  |  |  |  |  |
| No | 672 (20.3) | 0.520 | 303 (9.17) | 0.277 | 517 (15.6) | 0.206 |
| Yes | 107 (21.8) |  | 54 (11.1) |  | 90 (18.4) |  |
| **Maternal characteristics** |  |  |  |  |  |  |
| Maternal age (years) |  |  |  |  |  |  |
| 15–19 | 10.4 (1.72) | 0.481 | 11.3 (3.2) | 0.086 | 11.3 (3.15) | 0.086 |
| 20–29 | 258 (42.5) |  | 131 (36.5) |  | 131 (36.5) |  |
| 30–39 | 296 (48.7) |  | 192 (53.6) |  | 192 (53.6) |  |
| 40–49 | 42.9 (7.06) |  | 24 (6.7) |  | 24 (6.72) |  |
| Maternal education |  |  |  |  |  |  |
| No education | 111 (18.2) | <0.001 | 64.5 (18.0) | <0.001 | 64.5 (18.0) | <0.001 |
| Primary | 296 (48.7) |  | 165 (46.2) |  | 165 (46.2) |  |
| Secondary or higher | 201 (33.1) |  | 128 (35.7) |  | 128 (35.7) |  |
| Maternal employment |  |  |  |  |  |  |
| Not working | 269 (44.3) | 0.006 | 161 (44.9) | 0.019 | 161 (44.9) | 0.019 |
| Working | 338 (55.7) |  | 197 (55.1) |  | 197 (55.1) |  |
| Maternal marital |  |  |  |  |  |  |
| Not married | 45.8 (7.55) | 0.241 | 22.4 (6.3) | 0.945 | 22.4 (6.28) | 0.945 |
| Married | 561 (92.5) |  | 335 (93.7) |  | 335 (93.7) |  |
| Household socioeconomic status |  |  |  |  |  |  |
| Wealth index |  |  |  |  |  |  |
| Poorest | 214 (35.2) | <0.001 | 110 (30.7) | 0.043 | 110 (30.7) | 0.043 |
| Poorer | 131 (21.6) |  | 72.1 (20.2) |  | 72.1 (20.2) |  |
| Middle | 106 (17.5) |  | 54.4 (15.2) |  | 54.4 (15.2) |  |
| Richer | 108 (17.7) |  | 68.2 (19.1) |  | 68.2 (19.1) |  |
| Richest | 48.7 (8.01) |  | 53.1 (14.9) |  | 53.1 (14.9) |  |
| Household size |  |  |  |  |  |  |
| Small (≤3) | 69.6 (11.5) | 0.011 | 45.1 (12.6) | 0.162 | 45.1 (12.6) | 0.162 |
| Medium (4–6) | 442 (72.9) |  | 255 (71.3) |  | 255 (71.3) |  |
| Large (≥7) | 95.1 (15.7) |  | 57.7 (16.1) |  | 57.7 (16.1) |  |
| Housing quality index |  |  |  |  |  |  |
| Unimproved | 409 (67.5) | <0.001 | 220 (61.6) | 0.251 | 220 (61.6) | 0.251 |
| Improved | 198 (32.5) |  | 137 (38.4) |  | 137 (38.4) |  |
| Household crowding |  |  |  |  |  |  |
| Not crowded | 220 (36.3) | 0.001 | 143 (40.0) | 0.219 | 143 (40.0) | 0.219 |
| Crowded | 387 (63.7) |  | 215 (60.0) |  | 215 (60.0) |  |
| Agricultural land ownership |  |  |  |  |  |  |
| Small (1–5 ha) | 97.8 (16.1) | 0.937 | 57.3 (16.0) | 0.941 | 57.3 (16.0) | 0.941 |
| Medium (5–20 ha) | 147 (24.3) |  | 86.1 (24.1) |  | 86.1 (24.1) |  |
| Large (>20 ha) | 362 (59.6) |  | 214 (59.9) |  | 214 (59.9) |  |
| **Water, sanitation, and hygiene (WASH)** |  |  |  |  |  |  |
| Drinking water source |  |  |  |  |  |  |
| Improved | 382 (62.9) | <0.001 | 213 (59.7) | 0.274 | 213 (59.7) | 0.274 |
| Bottled | 126 (20.8) |  | 95 (26.6) |  | 95 (26.6) |  |
| Unimproved | 99.1 (16.3) |  | 49.3 (13.8) |  | 49.3 (13.8) |  |
| Household water treatment |  |  |  |  |  |  |
| No treatment | 211 (34.7) | 0.060 | 140 (39.2) | 0.004 | 140 (39.2) | 0.004 |
| Treated | 397 (65.3) |  | 217 (60.8) |  | 217 (60.8) |  |
| Sanitation facility type |  |  |  |  |  |  |
| Improved | 485 (79.9) | <0.001 | 297 (83.1) | 0.099 | 297 (83.1) | 0.099 |
| Unimproved | 12.4 (2.04) |  | 6.18 (1.7) |  | 6.18 (1.73) |  |
| Open defecation | 110 (18.1) |  | 54.3 (15.2) |  | 54.3 (15.2) |  |
| Handwashing facility availability |  |  |  |  |  |  |
| Not ready | 183 (30.1) | 0.002 | 87.3 (24.4) | 0.947 | 87.3 (24.4) | 0.947 |
| Ready (fixed place & water) | 424 (69.9) |  | 270 (75.6) |  | 270 (75.6) |  |
| Cooking fuel type |  |  |  |  |  |  |
| Clean | 247 (40.7) | <0.001 | 165 (46.2) | 0.088 | 165 (46.2) | 0.088 |
| Non-clean | 360 (59.3) |  | 192 (53.8) |  | 192 (53.8) |  |
| Household electricity access |  |  |  |  |  |  |
| No | 96.9 (16.0) | <0.001 | 42.7 (12.0) | 0.061 | 42.7 (12.0) | 0.061 |
| Yes | 510 (84.0) |  | 315 (88.0) |  | 315 (88.0) |  |
| Household refrigerator ownership |  |  |  |  |  |  |
| No | 517 (85.2) | <0.001 | 286 (80.0) | 0.004 | 286 (80.0) | 0.004 |
| Yes | 89.7 (14.8) |  | 71.4 (20.0) |  | 71.4 (20.0) |  |
| Household smoking |  |  |  |  |  |  |
| No | 358 (59.0) | 0.001 | 213 (59.5) | 0.019 | 213 (59.5) | 0.019 |
| Yes | 249 (41.0) |  | 145 (40.5) |  | 145 (40.5) |  |
| Health access and media exposure |  |  |  |  |  |  |
| Access time to facility |  |  |  |  |  |  |
| ≤15 min | 348 (57.4) | 0.061 | 207 (57.9) | 0.155 | 207 (57.9) | 0.155 |
| 16–30 min | 208 (34.3) |  | 118 (33.0) |  | 118 (33.0) |  |
| >30 min | 50.4 (8.3) |  | 32.7 (9.1) |  | 32.7 (9.14) |  |
| Health insurance coverage |  |  |  |  |  |  |
| No insurance | 455 (74.9) | 0.508 | 269 (75.1) | 0.663 | 269 (75.1) | 0.663 |
| Covered | 153 (25.1) |  | 89.1 (24.9) |  | 89.1 (24.9) |  |
| Media exposure |  |  |  |  |  |  |
| None | 359 (59.1) | 0.200 | 201 (56.2) | 0.319 | 201 (56.2) | 0.319 |
| One | 178 (29.4) |  | 116 (32.5) |  | 116 (32.5) |  |
| Two or more | 69.8 (11.5) |  | 40.2 (11.2) |  | 40.2 (11.2) |  |
| Mobile phone ownership |  |  |  |  |  |  |
| None | 138 (22.8) | <0.001 | 74.2 (20.8) | 0.019 | 74.2 (20.8) | 0.019 |
| Non-smartphone | 73.6 (12.1) |  | 39.7 (11.1) |  | 39.7 (11.1) |  |
| Smartphone | 395 (65.1) |  | 244 (68.1) |  | 244 (68.1) |  |
| Internet use |  |  |  |  |  |  |
| Never | 316 (52.0) | <0.001 | 168 (46.9) | 0.005 | 168 (46.9) | 0.005 |
| Weekly | 61.7 (10.2) |  | 44.4 (12.4) |  | 44.4 (12.4) |  |
| Daily | 230 (37.8) |  | 145 (40.7) |  | 145 (40.7) |  |
| Geographic variables |  |  |  |  |  |  |
| Residence |  |  |  |  |  |  |
| Urban | 169 (27.8) | <0.001 | 114 (31.9) | 0.105 | 114 (31.9) | 0.105 |
| Rural | 438 (72.2) |  | 244 (68.1) |  | 244 (68.1) |  |
| Geographic region |  |  |  |  |  |  |
| Phnom Penh | 45.2 (7.45) | 0.003 | 29.4 (8.2) | 0.132 | 29.4 (8.21) | 0.132 |
| Plain | 190 (31.2) |  | 116 (32.4) |  | 116 (32.4) |  |
| Tonle Sap | 235 (38.7) |  | 148 (41.3) |  | 148 (41.3) |  |
| Coastal | 33.9 (5.58) |  | 19.6 (5.5) |  | 19.6 (5.48) |  |
| Mountain | 103 (17.0) |  | 45 (12.6) |  | 45 (12.6) |  |

**Note:** (n) represents the weighted count and (%) weighted percentage. Design-based F-test p-values reported.

### Stunting

The prevalence of stunting increased significantly with child age, starting at 12.3% among infants (0–11 months) and peaking at 28.0% in children aged 12–23 months (p < 0.001). Boys had a higher prevalence of stunting (56.8%) compared to girls (43.2%). Although stunting rates were slightly higher among children with a recent history of diarrhea (24.4%) compared to those without (20.3%), this difference was not statistically significant. A sharp socioeconomic gradient was evident; stunting prevalence was more than four times higher in the poorest wealth quintile (35.2%) than in the richest (8.0%, p < 0.001). Similarly, children whose mothers had secondary or higher education had a lower prevalence (33.1%) than those whose mothers had no formal education (18.2%) or only primary education (48.7%, p < 0.001). Household and environmental conditions also played a major role: stunting was significantly more common among children living in unimproved housing (67.5%), crowded households (63.7%), and homes using non-clean cooking fuels (59.3%). Disparities were also observed regarding drinking water sources, sanitation, electricity, and access to technology (all p < 0.05).

### Wasting

Overall, the differences in wasting prevalence across characteristics were smaller than those for stunting. However, boys still showed a higher prevalence (58.2%) than girls (41.8%). While children with a recent fever or diarrhea had slightly higher rates of wasting, these associations were not statistically significant. Socioeconomic status remained a factor, with the poorest households contributing the largest share of cases (30.7%) and the richest households contributing the smallest (14.9%, p = 0.043). Higher proportions of wasting were also present among children of mothers with no education and those from households with limited access to refrigerators, internet, and treated water.

### Underweight

The patterns for underweight were very similar to those for wasting. Boys accounted for a higher proportion of cases (58.2%) than girls. There was no statistically significant link between recent illness (diarrhea or fever) and being underweight. Maternal education and household wealth were again key drivers: underweight was more prevalent among children of mothers with no education (18.0%) and those from the poorest wealth quintile (30.7%). Consistent with the other indicators, environmental factors like lack of water treatment and smoking exposure were associated with higher proportions of underweight children.

#### Multicollinearity

Multicollinearity among independent variables was assessed using an Ordinary Least Squares (OLS) regression model as a diagnostic proxy, followed by examination of Variance Inflation Factors (VIFs). This approach was applied to evaluate potential linear dependence among predictors included in the multivariable models.

The results indicate that multicollinearity is not a concern in the final specification. The overall mean VIF was 1.68, which is well below the commonly accepted threshold of 10 and also below the more conservative cut-off value of 5, indicating a low level of correlation among explanatory variables.

Most variables exhibited VIF values close to 1–2, suggesting minimal multicollinearity. The highest VIF was observed for the wealth index at 6.48, followed by cooking fuel type (2.54) and housing quality (1.93). Although these variables show moderate inter-correlation, all values remain below the critical threshold of 10, indicating that they do not pose a serious threat to the stability or interpretability of the regression estimates [17].

#### Goodness-of-Fit

The goodness-of-fit of the survey-weighted logistic regression models was assessed using the F-adjusted mean residual test. Overall, the results indicate that the models provide an adequate fit to the observed data.

For stunting, the model demonstrated strong fit (F = 0.43, p = 0.918), indicating no evidence of lack of fit and suggesting that the model appropriately captures the observed variation in chronic malnutrition. Similarly, the underweight model showed good fit (F = 0.87, p = 0.549), confirming adequate model specification.

The wasting model also showed acceptable fit (F = 0.64, p = 0.765), with no statistically significant evidence of model misspecification.

### Model Performance and Discriminatory Ability

The predictive performance of the multivariable logistic regression models for stunting, wasting, and underweight was evaluated using the area under the receiver operating characteristic (ROC) curve (AUC) (**Fig. 1**).

**Fig 1.**
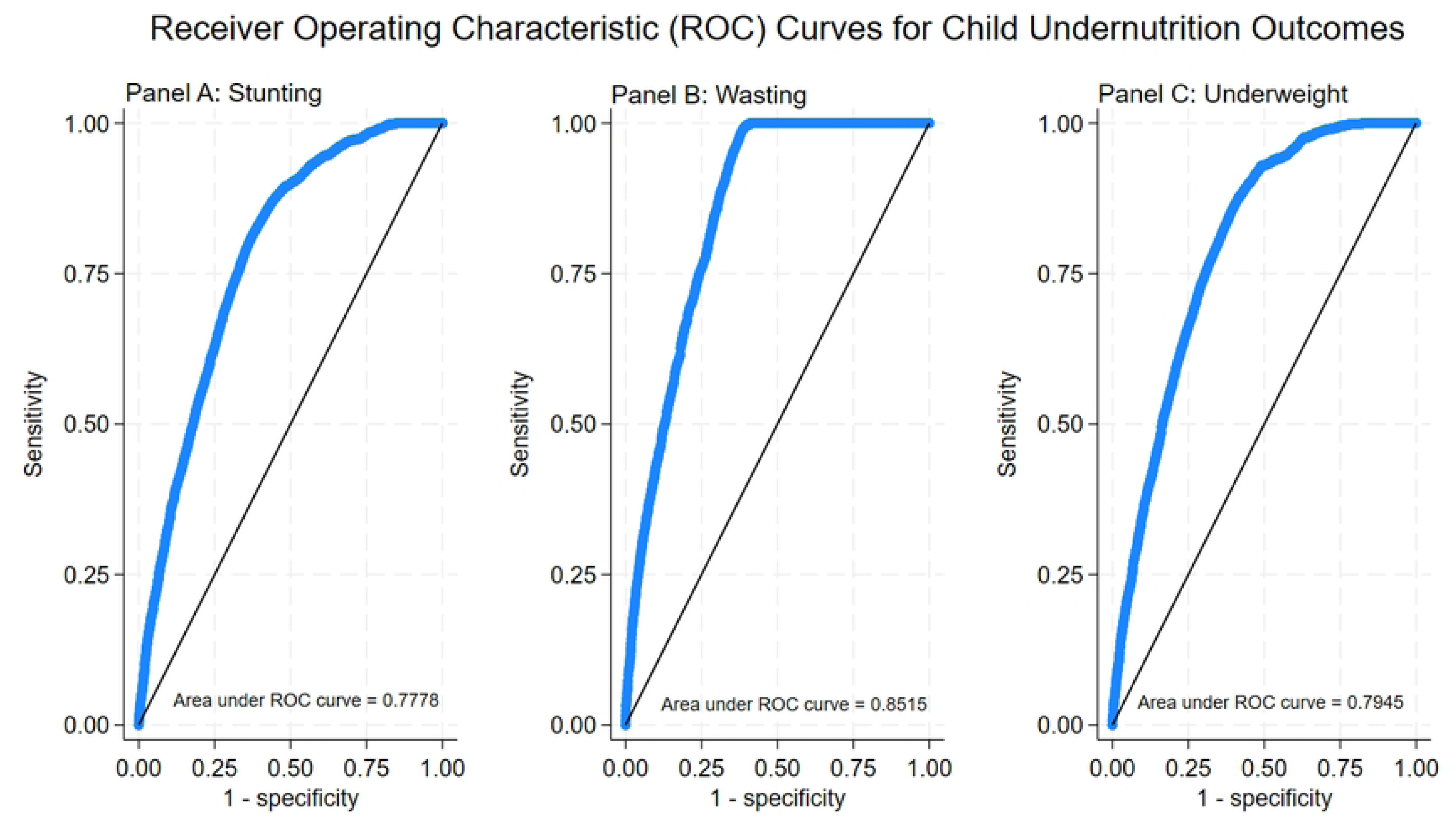
Receiver operating characteristic (ROC) curves for logistic regression models predicting (A) stunting, (B) wasting, and (C) underweight among children aged 0–59 months in Cambodia (CDHS 2021–2022).

Overall, all models demonstrated acceptable to good discriminatory ability. The model for wasting showed the best performance (AUC = 0.852, 95% CI: 0.837–0.866), indicating strong discrimination between wasted and non-wasted children. This was followed by underweight (AUC = 0.795, 95% CI: 0.779–0.810) and stunting (AUC = 0.778, 95% CI: 0.762–0.793), both of which demonstrated acceptable model performance.

### Adjusted Odds Ratios for Child Stunting, Wasting, and Underweight

The adjusted logistic regression model for stunting (**Table 3**) identified several significant determinants.

**Table 3.**
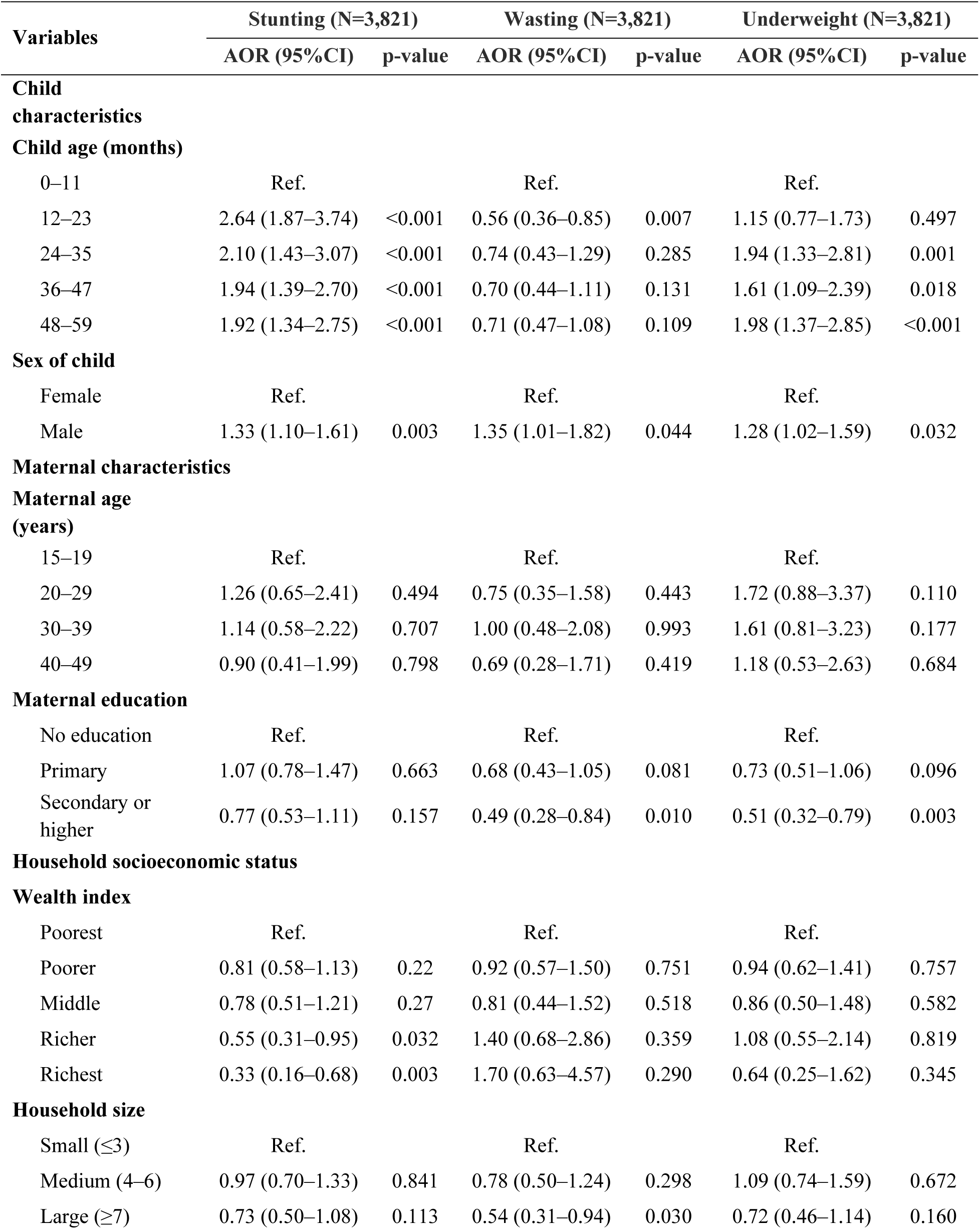
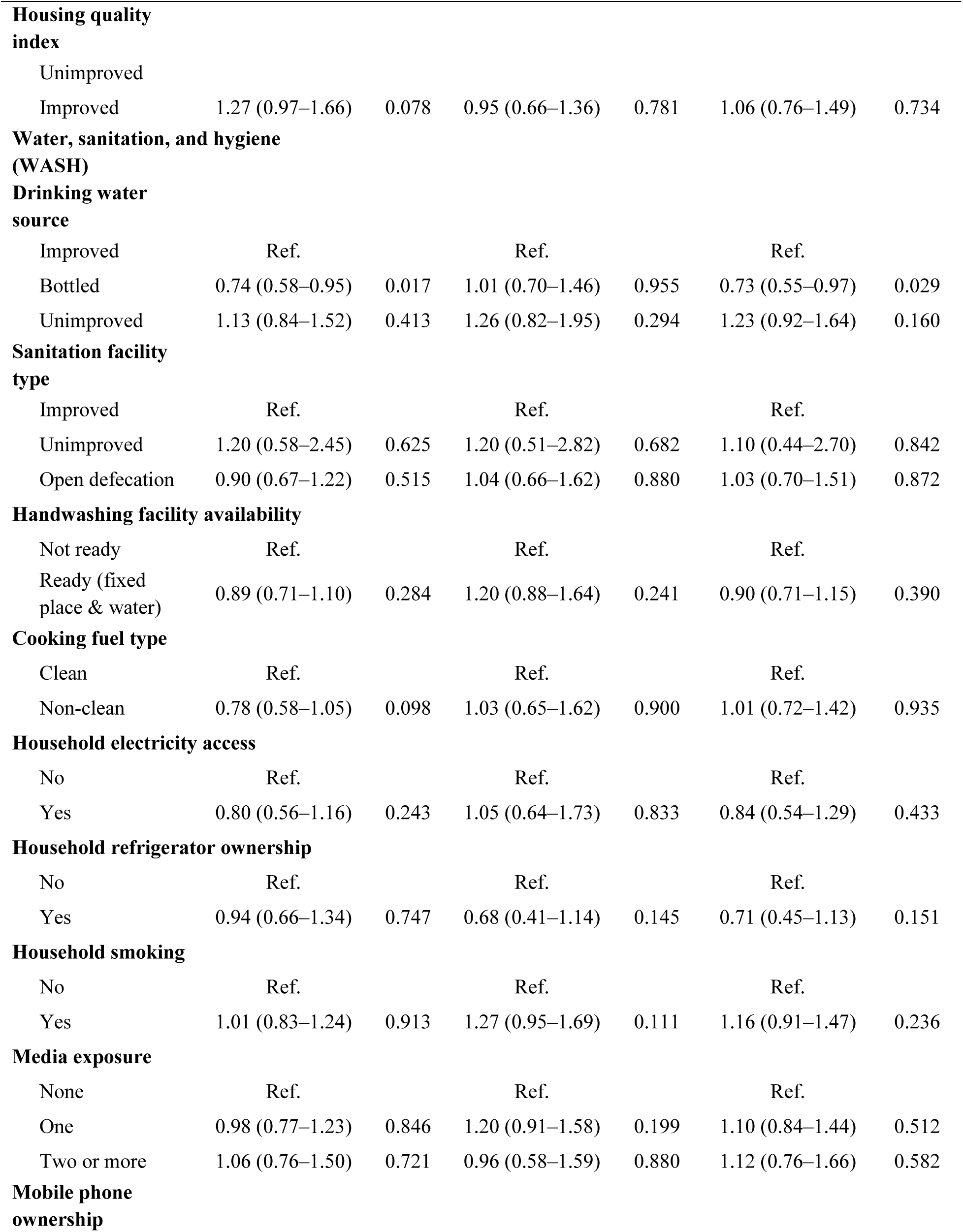
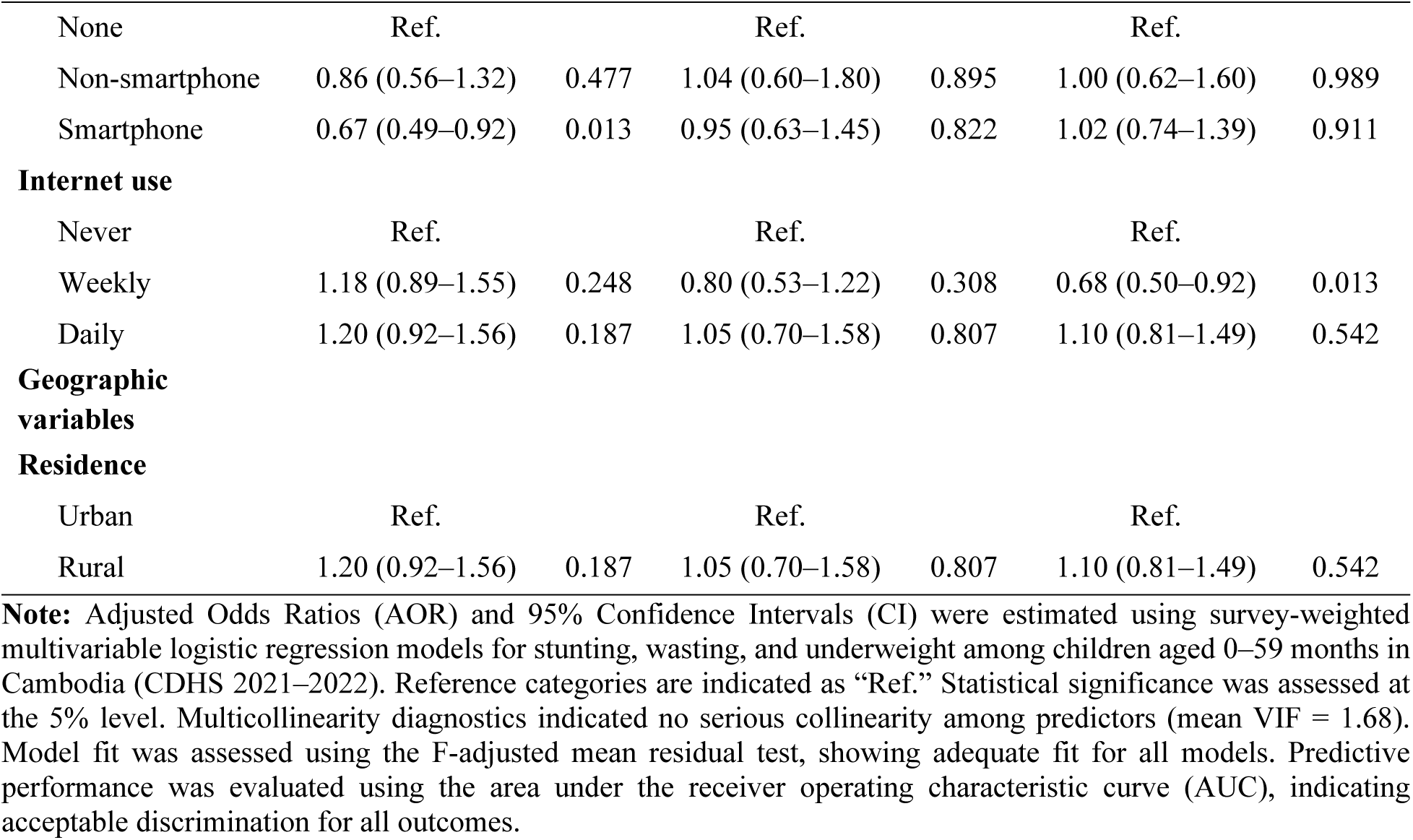
Adjusted Odds Ratios (AOR) and 95% Confidence Intervals for Factors Associated with Stunting, Wasting, and Underweight Among Children Under Five in Cambodia (CDHS 2021–2022)

### Factors Associated with Stunting

The multivariable analysis showed that child age was strongly associated with stunting. Compared with infants aged 0–11 months, the odds of stunting were significantly higher among children aged 12–23 months (AOR = 2.64, 95% CI: 1.87–3.74), 24–35 months (AOR = 2.10, 95% CI: 1.43– 3.07), 36–47 months (AOR = 1.94, 95% CI: 1.39–2.70), and 48–59 months (AOR = 1.92, 95% CI: 1.34–2.75). Male children had higher odds of stunting compared with females (AOR = 1.33, 95% CI: 1.10–1.61).

Household socioeconomic status showed a clear protective gradient, with reduced odds of stunting among children from richer (AOR = 0.55, 95% CI: 0.31–0.95) and richest households (AOR = 0.33, 95% CI: 0.16–0.68) compared with the poorest. Access to bottled drinking water was also associated with lower odds of stunting (AOR = 0.74, 95% CI: 0.58–0.95). In addition, children in households with smartphone ownership had reduced odds of stunting (AOR = 0.67, 95% CI: 0.49–0.92). These findings suggest that both biological vulnerability (age and sex) and structural determinants (wealth and digital access) jointly influence chronic malnutrition.

### Factors Associated with Wasting

Child age showed an inverse association with wasting. Children aged 12–23 months had lower odds of wasting compared with infants aged 0–11 months (AOR = 0.56, 95% CI: 0.36–0.85). No significant differences were observed for older age groups.

Male children had higher odds of wasting compared with females (AOR = 1.35, 95% CI: 1.01–1.82). Maternal education was an important protective factor; children of mothers with secondary or higher education had lower odds of wasting (AOR = 0.49, 95% CI: 0.28–0.84). Larger household size (≥7 members) was also associated with reduced odds of wasting (AOR = 0.54, 95% CI: 0.31–0.94). This pattern may reflect the more acute and short-term nature of wasting compared to stunting.

### Factors Associated with Underweight

Older age groups were consistently associated with higher odds of underweight. Compared with infants, children aged 24–35 months (AOR = 1.94, 95% CI: 1.33–2.81), 36–47 months (AOR = 1.61, 95% CI: 1.09–2.39), and 48–59 months (AOR = 1.98, 95% CI: 1.37–2.85) were significantly more likely to be underweight.

Male children had higher odds of underweight compared with females (AOR = 1.28, 95% CI: 1.02–1.59). Maternal education was strongly protective; children of mothers with secondary or higher education had lower odds of underweight (AOR = 0.51, 95% CI: 0.32–0.79). Access to bottled drinking water was associated with reduced odds of underweight (AOR = 0.73, 95% CI: 0.55–0.97). Weekly internet use was also protective against underweight (AOR = 0.68, 95% CI: 0.50–0.92). Underweight appears to capture both chronic and acute nutritional deficits, explaining its overlap with determinants identified for both stunting and wasting.

### Interaction effects on child undernutrition

Interaction analyses were conducted to assess whether the associations between key socioeconomic factors and child nutritional outcomes varied across subgroups. The models included interaction terms between child age and sex, household wealth and place of residence, maternal education and wealth.

No statistically significant interactions were observed between child age and sex or between household wealth and place of residence for any of the undernutrition outcomes (stunting, wasting, or underweight), suggesting that these associations were consistent across these subgroups. In contrast, a statistically significant interaction between maternal education and household wealth was consistently observed across all three outcomes. This finding indicates a synergistic protective effect, whereby the combined advantage of higher maternal education and higher household wealth is associated with a substantially lower risk of undernutrition. The lowest predicted risk was observed among children whose mothers had secondary or higher education and who resided in the richest households (**Fig 2**, **Fig 3**, **Fig 4**). This finding underscores the importance of considering combined socioeconomic advantages rather than isolated determinants when designing interventions.

**Fig 2.**
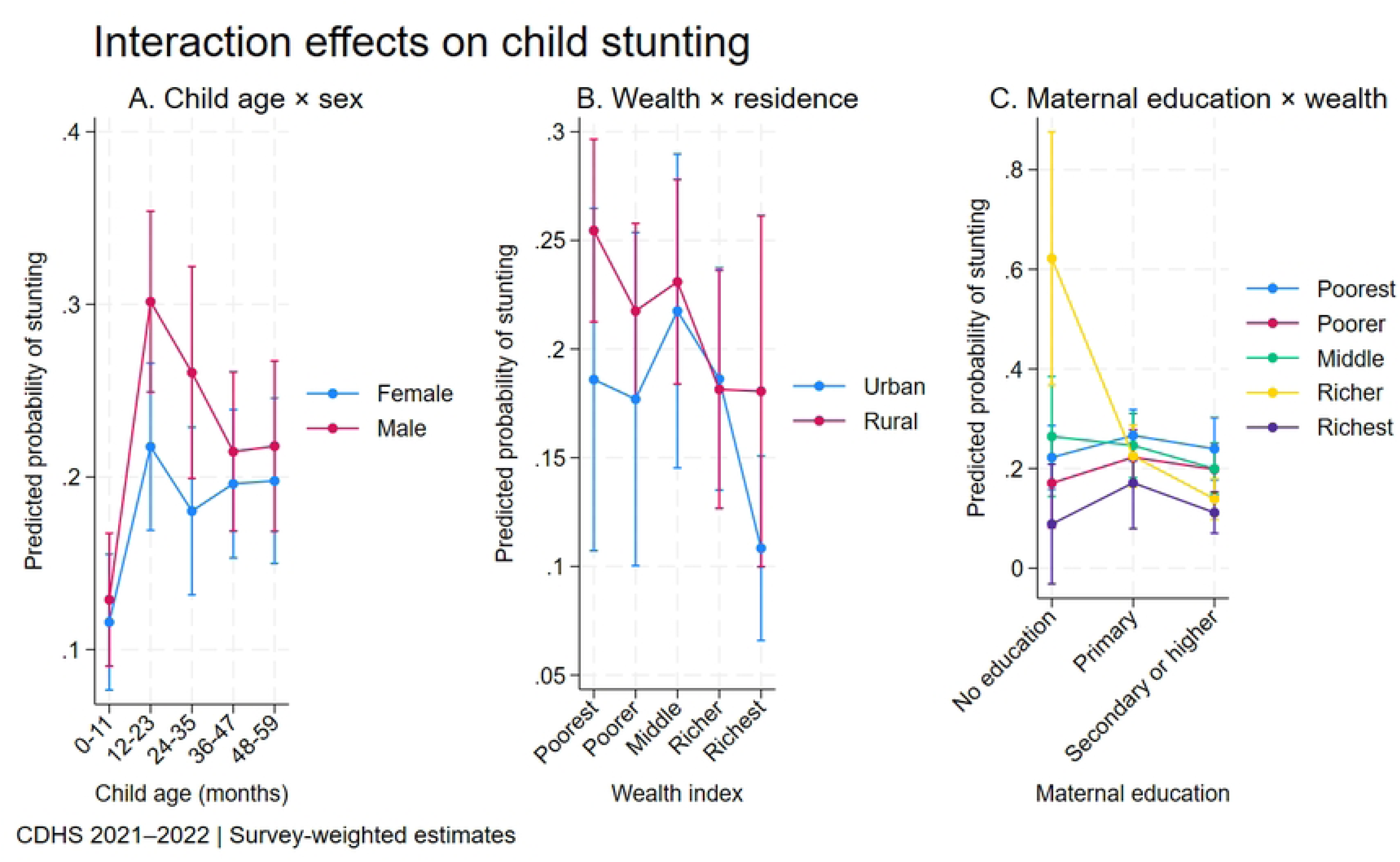
Interaction effects of socioeconomic factors on child stunting among children aged 0–59 months in Cambodia (CDHS 2021–2022). (A) Child age × sex; (B) Household wealth × residence; (C) Maternal education × household wealth.

**Fig 3.**
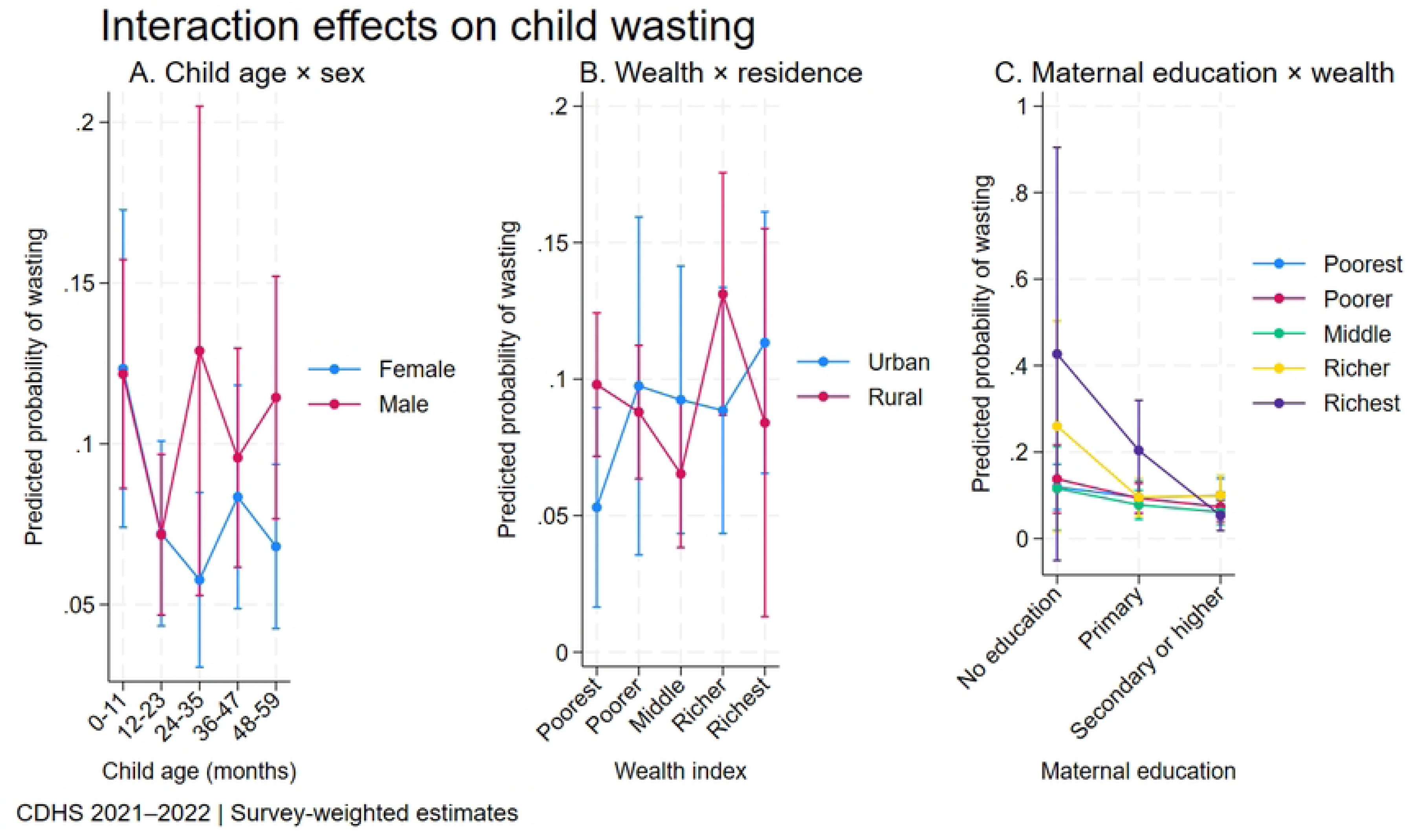
Interaction effects of socioeconomic factors on child wasting among children aged 0–59 months in Cambodia (CDHS 2021–2022). (A) Child age × sex; (B) Household wealth × residence; (C) Maternal education × household wealth.

**Fig 4.**
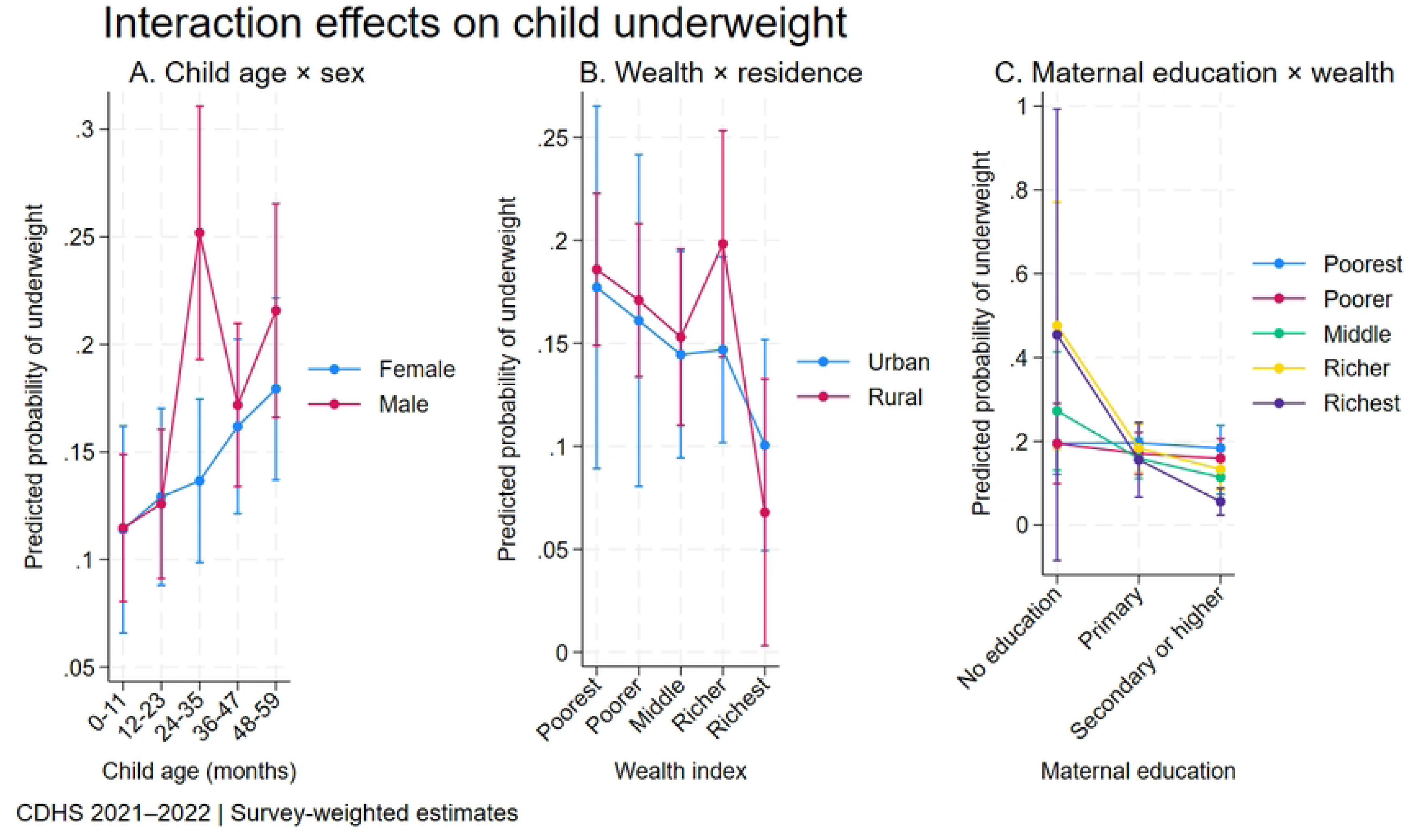
Interaction effects of socioeconomic factors on child underweight among children aged 0–59 months in Cambodia (CDHS 2021–2022). (A) Child age × sex; (B) Household wealth × residence; (C) Maternal education × household wealth.

## Discussion

This nationally representative study provides updated evidence on the prevalence and determinants of undernutrition among children aged 0–59 months in Cambodia. The findings reinforce the persistence of undernutrition as a multifactorial and inequality-driven public health challenge in Cambodia.

While the overall prevalence of stunting (20.4%), wasting (9.4%), and underweight (15.9%) reflects progress over the last two decades, these figures remain above the WHO thresholds for low public health concern [19]. Similar burdens in other lower-middle-income countries in Southeast Asia suggest shared structural and nutritional vulnerabilities across the region [20].

A critical finding is the strong age gradient in stunting, which peaks among children aged 12–23 months. This period represents a biologically sensitive window where inadequate dietary intake and increased exposure to infections can have irreversible effects on growth and development. CDHS 2021–2022 data indicate that dietary diversity and minimum acceptable diets remain suboptimal in Cambodia, particularly for children in poorer households [6]. Suboptimal feeding practices, coupled with repeated infections, likely drive growth faltering during this “first 1,000 days” window—a priority area for Cambodia’s National Nutrition Strategy 2023–2030 [12].

Socioeconomic status emerged as a dominant driver for all forms of undernutrition. The consistently higher prevalence among the poorest households reflects systemic inequities in income, food security, and healthcare access. This highlights that economic growth alone is insufficient without equitable distribution and targeted social protection mechanisms. While the IDPoor targeting system and Social Protection Policy Framework 2016–2025 have aimed to bridge these gaps through cash transfers, the persistence of these inequalities suggests that current coverage is not yet sufficient to offset structural deprivation [21]. This pattern is consistent with previous studies in Cambodia, including research on childhood anemia, which has shown significantly higher risks among children from poorer households [8]. Similarly, studies on ARI and diarrhea have identified household wealth and maternal education as key protective factors, reinforcing the central role of socioeconomic inequality across multiple child health outcomes [9, 10].

Maternal education was found to be strongly protective, particularly against wasting and underweight. This aligns with global evidence that educated mothers are often better equipped to utilize health services and implement effective childcare practices [22]. This underscores the importance of the Education Strategic Plan (2019–2023) as a long-term health investment [23]. Evidence from ARI and diarrhea studies in Cambodia similarly highlights maternal education as a critical determinant, influencing health-seeking behavior, hygiene practices, and the ability to prevent and manage childhood illnesses [9, 10].

Environmental conditions remain major determinants, with higher undernutrition rates found in unimproved or crowded housing and areas lacking clean water and sanitation. These results reinforce the necessity of the National Action Plan for Rural WASH (2019–2023) [24]. These findings are strongly supported by prior research on childhood diarrhea and ARI, which has demonstrated that poor WASH conditions and exposure to environmental risk factors—such as unsafe water, inadequate sanitation, and household smoking—significantly increase child morbidity. Such repeated exposure to infections may indirectly contribute to undernutrition through environmental enteropathy, leading to impaired nutrient absorption and increased metabolic demands [9, 10].

Interestingly, digital access (smartphones and internet) also appeared protective, suggesting that digital connectivity may enhance maternal knowledge and health-seeking behavior. This supports Cambodia’s digital transformation agenda and the potential for emerging mHealth initiatives [25–27].

Notably, recent childhood illnesses like diarrhea and fever were not significantly associated with undernutrition in the multivariable models. This may be due to the cross-sectional nature of the data [6], but it also supports the WHO conceptual framework: long-term nutritional status is driven more by chronic structural factors—such as poverty and sanitation—than by isolated acute episodes [4, 20]. This finding may also reflect the broader epidemiological transition observed in Cambodia, where substantial reductions in infectious diseases such as ARI and diarrhea have been achieved over time. As a result, chronic determinants—particularly socioeconomic and environmental conditions—may now play a more dominant role in shaping child nutritional outcomes [9, 10].

Interaction analysis revealed that the protective effect of maternal education is stronger in wealthier households, where mothers have the resources to act on their health knowledge. In contrast, structural poverty in the poorest households may limit the benefits of education. This reflects “capability constraints,” where knowledge alone is insufficient without the economic and environmental resources required to translate it into improved child nutrition. This indicates that shared environmental and health system factors at the local level significantly influence outcomes, justifying the need for geographically targeted interventions [4, 20].

Taken together, evidence from this study and prior research on anemia, ARI, and diarrhea suggests that Cambodia is undergoing a partial epidemiological transition in child health. While infectious diseases have declined significantly, nutrition-related conditions—particularly those linked to chronic socioeconomic disadvantage—remain persistent [9, 10]. This underscores the need for integrated, multisectoral strategies that simultaneously address nutrition, poverty, environmental conditions, and access to health information and services.

### Public Health Implications

The findings have important implications for Cambodia’s national nutrition and development agenda.

First, the high burden of stunting among children aged 12–23 months underscores the need to strengthen complementary feeding interventions during the critical 6–23 months window. The National Nutrition Strategy 2023–2030 should further prioritize scaling up infant and young child feeding (IYCF) programs, including dietary diversity promotion, breastfeeding counseling, and community-based nutrition support through health centers and village health support groups (MoH Cambodia, 2023) [12]. CDHS IYCF data further highlight the need to improve minimum acceptable diet coverage among young children, particularly in rural and poor households (NIS et al., 2023) [6].

Second, the strong socioeconomic gradient in undernutrition highlights the importance of strengthening equity-focused social protection systems. Expanding and improving the IDPoor system is essential to ensure effective targeting of vulnerable households for cash transfers, food assistance, and maternal-child health services (RGC, 2017) [21].

Third, maternal education remains a key long-term determinant. Sustained investment in girls’ education should be considered a core nutrition-sensitive strategy. In the short term, community-based nutrition education delivered through health centers and outreach workers should be expanded.

Fourth, environmental determinants highlight the need for cross-sectoral collaboration between the Ministry of Health, Ministry of Rural Development, and related sectors. Improving access to clean water, sanitation, clean cooking energy, and adequate housing should be fully integrated into national child health strategies (MoRD, 2019) [24].

Fifth, digital health interventions represent an emerging opportunity. Expansion of mHealth and digital nutrition communication platforms could improve maternal knowledge and service uptake, supporting Cambodia’s Digital Health Transformation Strategy (2021–2035) (MoH Cambodia, 2021) [26, 27].

Finally, these findings strongly support Cambodia’s progress toward Sustainable Development Goal 2 (Zero Hunger) and related SDGs on health, education, and inequality reduction (UN, 2015) [28]. Achieving these goals will require integrated multisectoral action addressing both immediate and structural determinants of malnutrition.

### Strengths and limitations

This study has several strengths. It uses nationally representative CDHS 2021–2022 data with appropriate survey weighting, ensuring strong external validity. The study includes three key anthropometric indicators (stunting, wasting, underweight), allowing a comprehensive assessment of both chronic and acute malnutrition. A wide range of determinants across child, maternal, household, environmental, and geographic domains were included.

Methodologically, the study applied survey-weighted multivariable logistic regression, multicollinearity diagnostics, interaction analysis, and model performance evaluation using ROC curves and goodness-of-fit tests, strengthening robustness.

However, limitations should be acknowledged. The cross-sectional design limits causal inference. Key dietary variables (including detailed IYCF indicators for 6–23 months such as dietary diversity and minimum acceptable diet) were not included in multivariable models, although they are available in CDHS. Self-reported illness data may be subject to recall bias. Residual confounding from unmeasured factors such as birth weight and maternal nutritional status may also remain.

## Conclusion

This study provides robust national evidence on child undernutrition in Cambodia using CDHS 2021–2022 data. Despite progress in national development, undernutrition remains a significant and unequal public health burden.

Children from poorer households, those with less educated mothers, and those living in disadvantaged environmental conditions bear the highest burden of stunting, wasting, and underweight. The critical vulnerability during the 6–23 months feeding window highlights the importance of improving infant and young child feeding practices.

Reducing child undernutrition in Cambodia will require sustained multisectoral action aligned with the National Nutrition Strategy 2023–2030, Social Protection Policy Framework, and WASH and Rural Development strategies. Strengthening maternal education, improving household environments, enhancing targeting systems, and expanding digital health interventions are essential pathways toward accelerating progress.

These integrated efforts are essential for Cambodia to achieve Sustainable Development Goal 2: Zero Hunger and ensure equitable child health and nutrition outcomes.

## Data Availability

The data used in this study are publicly available from the Demographic and Health Surveys (DHS) Program. The 2021–22 Cambodia Demographic and Health Survey (CDHS) dataset can be accessed upon reasonable request and approval from https://dhsprogram.com. The authors did not have any special access privileges to the data.

## Acknowledgments

The authors would like to thank DHS-ICF for approving the data used in this paper.

## Abbreviations

AOR: Adjusted Odds Ratio
AUC: Area Under the Curve
CDHS: Cambodia Demographic and Health Survey
CI: Confidence Interval
DHS: Demographic and Health Survey
GDP: Gross Domestic Product
IYCF: Infant and Young Child Feeding
LMICs: Low- and Middle-Income Countries
MoH: Ministry of Health
mHealth: Mobile Health
NIS: National Institute of Statistics
OLS: Ordinary Least Squares
ROC: Receiver Operating Characteristic
SDG: Sustainable Development Goal
VIF: Variance Inflation Factor
WASH: Water, Sanitation, and Hygiene
WHO: World Health Organization

